# Real-time Continuous Measurement of Lactate through a Minimally-invasive Microneedle Biosensor: a Phase I Clinical Study

**DOI:** 10.1101/2021.08.23.21262407

**Authors:** DK Ming, S Jangam, SAN Gowers, R Wilson, DME Freeman, MG Boutelle, AEG Cass, D O’Hare, AH Holmes

## Abstract

**Introduction:** Determination of blood lactate levels supports decision-making in a range of medical conditions. Invasive blood-sampling and laboratory access are often required, and measurements provide a static profile at each instance. We conducted a Phase I clinical study validating performance of a microneedle patch for minimally-invasive, continuous lactate measurement in healthy volunteers.

**Methods:** Five healthy adult participants wore a solid microneedle biosensor on their forearms and undertook aerobic exercise for 30 minutes. The microneedle biosensor quantifies lactate concentrations in interstitial fluid (ISF) within the dermis continuously and in real-time. Outputs were captured as sensor current and compared with lactate concentrations from venous blood and microdialysis.

**Results:** The biosensor was well-tolerated. Participants generated a median peak venous lactate of 9.25 mmol/L (Interquartile range, 6.73 to 10.71). Microdialysate concentrations of lactate closely correlated with blood. Microneedle biosensor current followed venous lactate concentrations and dynamics, with good agreement seen in all participants. There was an estimated lag-time of 5 minutes (IQR -4 to 11 minutes) between microneedle and blood lactate measurements.

**Conclusion:** This study provides first-in-human data on use of a minimally-invasive microneedle biosensor for continuous lactate measurement, providing dynamic monitoring. The platform offers distinct advantages to frequent blood sampling in a wide range of clinical settings, especially where access to laboratory services is limited or blood sampling is infeasible.

## Introduction

Raised lactate concentrations in blood are associated with all-cause mortality in hospitalised patients^1^. The dynamics of lactate change over time and higher rates of early clearance are also associated with favourable responses to therapy and improved clinical outcomes^2^. Lactate levels in humans result from the physiological interplay between tissue perfusion, hepatic and renal clearance, tissue hypoxia and the rate of glycolysis^3^. Use of lactate as a biomarker to guide medical therapy and risk-stratification has been extensively validated, and supports management of infections such as sepsis^4^, malaria^5^ and dengue^6^, as well as in trauma^7^, acute heart failure^8^, interoperative optimisation^9^, and exercise training^10^.

In acute clinical settings, blood lactate concentrations are commonly quantified using laboratory analysers. Availability of bedside point-of-care measurements through blood gas analysers and capillary lactate devices further improves access to testing by reducing turnaround time. However, contemporary measurement methods all require blood sampling: venous puncture or expert arterial puncture are uncomfortable and can lead to complications, and the poor concordance of capillary lactate with whole blood restricts its use in clinical settings^11^. Venesection poses particular challenges in special populations such as neonates or children, and the timely analysis of blood samples is often not feasible in healthcare settings with limited access to laboratory services^12^. Where frequent measurements of lactate are clinically indicated– repeated sampling may be facilitated through placement of an arterial catheter and implantable intravenous continuous sensors have also been proposed^13^. These invasive interventions are not possible beyond critical care settings and therefore preclude the role of frequent lactate monitoring as an adjunct to decision-making in prehospital, community healthcare or resource-limited settings.

Recent developments have supported minimally-invasive sampling of a range of bodily fluids. Analysis of interstitial fluid (ISF) in particular is promising – as the primary constituent of extracellular fluid, the compartment exists in dynamic equilibrium with plasma^14^. Relationships between blood and ISF lactate in pathology are complex, and have been described in hospitalised patients using microdialysis, an invasive ISF sampling technique^15,16^. In a cohort of patients with sepsis in intensive care, changes in ISF lactate levels preceded changes in blood lactate, suggesting the former could serve as a sensitive and early marker of pathology at the local tissue level^16^.

Measurement of substrates within the ISF is enabled through use of minimally-invasive platforms such as the microneedle biosensor^18^. The small device consists of a plastic base with arrays containing 1 mm protrusions, with each array acting as individual biosensors. When the microneedle biosensor is placed on skin surface, these protrusions are in continuous, direct contact with ISF within the viable epidermis and dermis. The electrochemical detection of lactate is mediated through an enzyme-based sensing biocompatible hydrogel layer (see Figure 1). An electrical current at the microneedle surface is measured and results can be displayed in real-time. As the microneedle protrusions in the skin lie superficial to the nerve layer, pain and discomfort is also minimised^19^.

**Figure 1.**
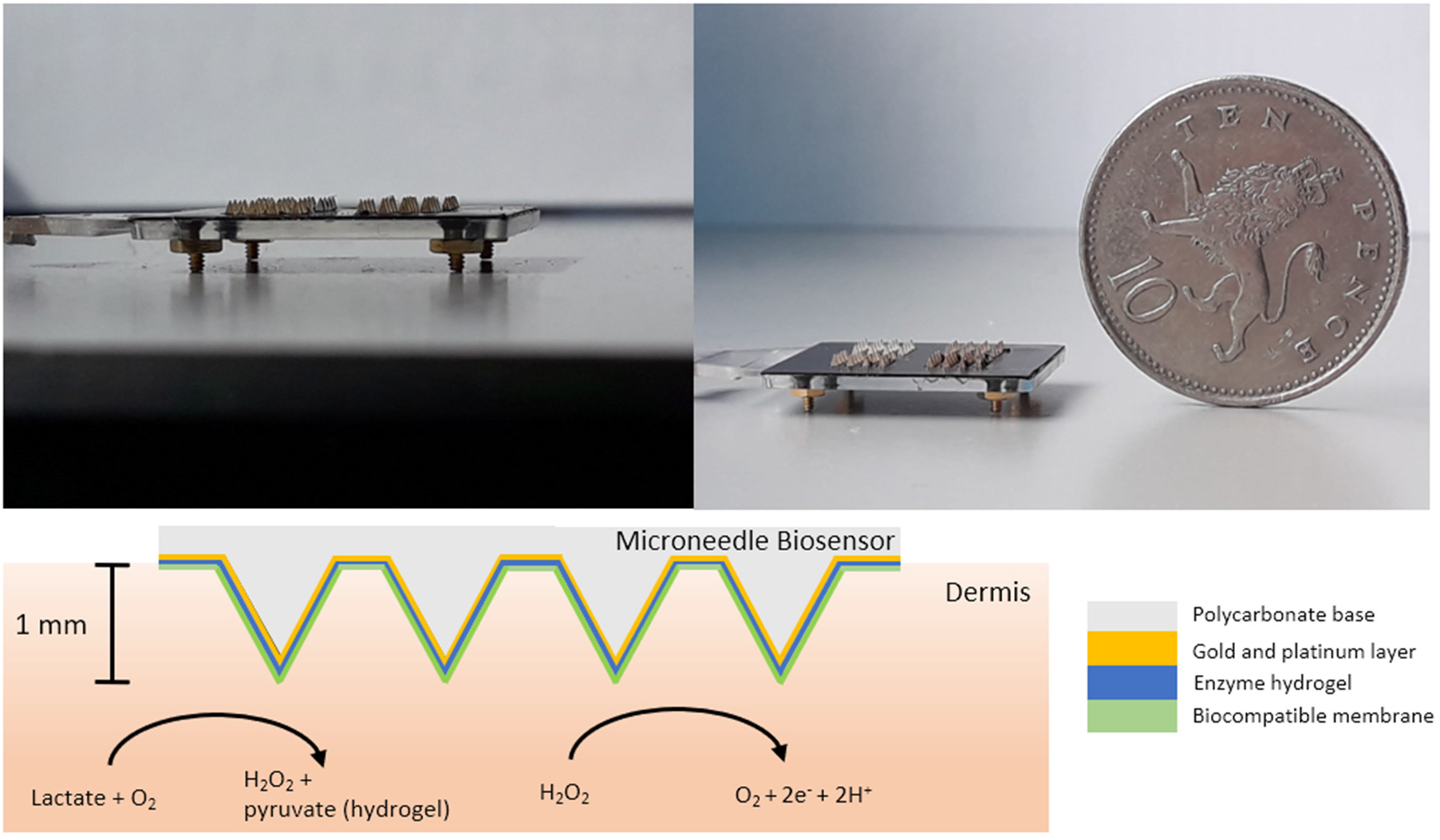
Top left and right: the microneedle biosensor measures less than 2 × 2 cm and consists of small 1 mm protrusions which penetrate the stratum corneum in the epidermis to come into direct contact with tissue interstitial fluid. Bottom: lactate in the interstitial fluid is converted to pyruvate and hydrogen peroxide, with the latter being oxidised at the biosensor electrode surface, which is held at +0.7 V vs Ag|AgCl reference electrode. The resulting current to ISF lactate concentration.

Clinical studies using the microneedle platform have demonstrated good performance and comfort in prolonged usage for up to 24 hours in glucose monitoring^20^ and penicillin monitoring^21^; findings from public and patient engagement events also support its acceptability and role in clinical monitoring when compared with blood sampling^22^.

We hypothesise that a minimally-invasive continuous lactate biosensor built on the microneedle platform could offer distinct patient and operational benefits resulting in improved clinical management in the healthcare setting: real-time continuous measurements are likely to directly inform clinical decision-making. We therefore conducted a first-in-human Phase I clinical study evaluating the performance of the microneedle-based lactate biosensor in healthy volunteers. Aerobic exercise was used as a proxy means of increasing body lactate concentrations. We determined ISF lactate concentrations using the microneedle biosensor and comparison with lactate levels obtained in venous blood. In order to characterise the relationship between venous and ISF lactate in exercise, microdialysis was used to provide a reference measurement.

## Methods

### Study design

This was a Phase I proof-of-concept clinical device study in healthy volunteers. The objective was evaluation of microneedle biosensors in measuring continuous interstitial fluid lactate in real-time before, during and after a short period of moderate aerobic exercise.

The study protocol was reviewed and approved by London - Bloomsbury Research Ethics Committee (20/LO/0364) and registered on Clinicaltrials.gov (NCT04238611).

The study was sponsored by Imperial College London and conducted at the National Institute of Health Research/Wellcome Trust Imperial Clinical Research Facility (Imperial College London, UK). All researchers underwent Good Clinical Practice training and procedures conducted in accordance with the 1964 Declaration of Helsinki and later amendments.

The approved study protocol is presented in the appendix.

### Participants

Between 16^th^ February 2021 and 10^th^ July 2021, participants were identified through recruitment posters placed around Imperial College London advertising the study. Male and female adult (18 years or older) healthy volunteers with no significant past medical history, who exercised regularly at moderate intensity for at least 30 minutes twice a week were eligible for inclusion. Exclusion criteria included any active inflammatory condition or infection of the skin, hypersensitivity to microneedle components or dressings, the presence of implantable electronic devices such as pacemakers, current pregnancy, active medication use, and active symptoms consistent with, or contact with anyone with COVID-19.

If participants responded to the advertisement, they were provided with a participant information leaflet in writing by email, followed by in-person screening on the day of the visit by the study physician.

### Sensor design and fabrication

The microneedle platform utilised in the study has been described previously^23^. Briefly, the microneedle array (Torr Scientific, UK) is made from a polycarbonate base which measures 2 × 2 cm and consists of 3 independent working electrode arrays, each consisting of 16 microneedles, and a Ag/AgCl/NaCl reference electrode on a fourth array. A layer of platinum black was applied onto the working electrode surfaces followed by deposition of a biocompatible enzyme-hydrogel layer, consisting of *lactate oxidase* enzyme from *Aerococcus viridans* (Sekisui Diagnostics, Japan), 2% glycerol, 3% w/v bovine serum albumin and 4% polyethylene glycol. Lactate generated in ISF is converted into pyruvate by the embedded enzyme within the hydrogel/skin interface, generating hydrogen peroxide proportionate to lactate concentration. The resulting hydrogen peroxide change is then amperometrically oxidised leading to a current at the platinum electrode surface measured by a potentiostat at the bedside. An inert membrane consisting of 1-5% Nafion (Sigma Aldrich, USA) was applied onto the electrode surface in multiple layers by nebulisation in order to extend dynamic range of detection. Single-use biosensors were fabricated up to a week ahead of the clinical study and kept refrigerated at 4°C until the day of use. Microneedle biosensors were calibrated against L-lactate standards before, and after clinical testing to confirm function.

### Procedures

All participants provided written informed consent and were assigned a study number. Procedures were performed to comply with COVID-19 precautions including the use of appropriate personal protective equipment throughout the study.

Lactate microneedle biosensors were placed on inner forearm skin surface prepared with 2% chlorhexidine gluconate / 70% isopropyl alcohol and applied using firm thumb pressure for 60 seconds. The biosensor was secured in place with an elasticated strap and had a wired connection to the potentiostat (CHI instruments, USA) with real-time microneedle current displayed on a laptop.

The microneedle biosensor was left in-situ on the forearm 60 minutes for stabilisation. The participant was then asked to cycle on an exercise bicycle (Ergoselect 200, Ergoline Germany) at 60 rpm at power increments of 35 W up to a maximum of 210 W, for a total of 30 minutes according to the protocol. The maximum exercise intensity reached was adjusted throughout the study according to participant ability and preference, but ensuring moderate activity was achieved. Power output values were recorded from the exercise bike. A rest period of 30 minutes with cessation of all exercise followed. Throughout the study period both forearms remained stationary on the handlebars. At the end of the rest period, the biosensor was removed and participants were given a questionnaire and photographs of the forearm obtained.

### Blood sampling

Venous lactate was sampled at regular 5-minute intervals over 60 minutes from an indwelling cannula in the contralateral arm to the biosensor using fluoride/oxalate vacutainer blood bottles (BD, USA). Samples were stored at 4°C and processed within 12 hours of collection at a UKAS-accredited laboratory located at North West London Pathology, Hammersmith Hospital UK. All samples were processed on the Architect Ci8200 analyser platform (Abbott, USA) through a colorimetric assay. All blood samples were discarded after 72 hours of analysis in line with local policy.

### Microdialysis

Microdialysis is an invasive method to sample and measure substrates in the interstitial fluid. A 63 microdialysis catheter (M Dialysis, Sweden) was inserted into the subcutaneous layer of the forearm using sterile technique. Topical anaesthesia (EMLA 5%) was applied on the skin surface prior to insertion. The inserted catheter was perfused with T1 sterile perfusion liquid (M Dialysis, Sweden) at a rate of 2 μL/minute allowing analytes, including lactate, in the subcutaneous ISF to diffuse into the catheter for downstream analysis. The catheter outflow was connected to a microfluidic chip housing an online lactate sensor developed in-house at Imperial College London^24^, connected to a portable potentiostat, which detected lactate continuously in real time. The microdialysis probe was inserted during 60 minutes of stabilisation, and continued until the end of the study period.

### Data processing and analysis

A robust LOWESS (locally weighted scatterplot smoothing) filter with a 60-second window was applied to raw microneedle and microdialysis data to remove outliers, motion artefact and attenuate noise. Venous lactate was modelled to change in a linear fashion based on physiological assumptions with additional data points interpolated at 1-minute intervals between blood measurements every 5 minutes. In order to calibrate individual sensors, we fitted individual standard curves using original venous lactate data against biosensor current for the lactate generation phase and clearance phase separately. The use of this technique for biosensor calibration is established in other settings such as continuous glucose monitoring ^25^.

We analysed the relationship between biosensor current and venous lactate dynamics, and ability of the biosensor to detect relative change in lactate by normalising biosensor current and venous lactate data to a range between 0 and 1. The mean rate of ISF and venous lactate change against time was then calculated using a 5-minute rolling window method and correlation described using Pearson’s coefficient. The time lag between biosensor and venous lactate was estimated at their respective points of inflection in the model when ISF and venous lactate concentrations were at their highest. All analyses and visualisations were done in Python 3.7 using the *pandas, scipy* and *matplotlib* libraries.

## Results

Five participants were enrolled into the study between 19^th^ May 2021 and 13^th^ July 2021 and assigned study numbers. One participant consented to undergoing microdialysis. The median age was 32 years (Interquartile range, IQR 27-33), and one (20%) participant was female. All participants completed 30 duration minutes of active exercise with a median power output of 113 W (IQR 93-130 W). The median baseline pre-exercise venous lactate was 1.40 mmol/L (IQR 1.23-1.52), peak venous lactate was 9.25 mmol/L (IQR 6.73 – 10.71), and venous lactate at the end of the rest period was 2.41 mmol/L (IQR 2.06-2.90). There were no adverse events reported during the study. Characteristics of the participants are shown in table 1.

**Table 1.**
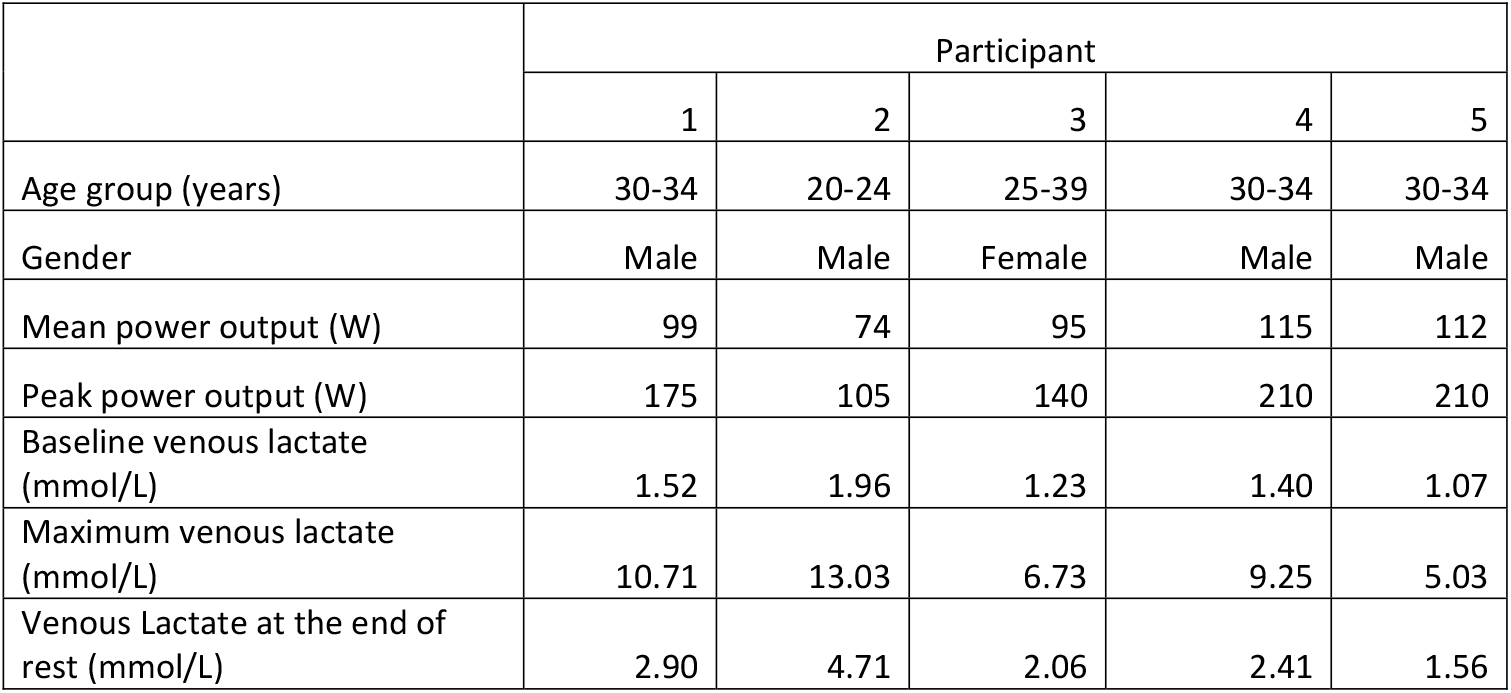
Characteristics of the 5 participants enrolled. All participants completed 30 minutes of active exercise with varying power outputs

|  | Participant |  |  |  |  |
| --- | --- | --- | --- | --- | --- |
|  | 1 | 2 | 3 | 4 | 5 |
| Age group (years) | 30-34 | 20-24 | 25-39 | 30-34 | 30-34 |
| Gender | Male | Male | Female | Male | Male |
| Mean power output (W) | 99 | 74 | 95 | 115 | 112 |
| Peak power output (W) | 175 | 105 | 140 | 210 | 210 |
| Baseline venous lactate (mmol/L) | 1.52 | 1.96 | 1.23 | 1.40 | 1.07 |
| Maximum venous lactate (mmol/L) | 10.71 | 13.03 | 6.73 | 9.25 | 5.03 |
| Venous Lactate at the end of rest (mmol/L) | 2.90 | 4.71 | 2.06 | 2.41 | 1.56 |

Microneedle biosensor current was plotted against time alongside venous lactate for each participant and shown in Figure 2. There was a rise and fall in venous blood lactate during the exercise (0-30 minutes) and rest (30-60 minutes) phases respectively. Continuous microneedle biosensor current followed venous concentrations closely over time. For participants 1-3, significant increases in biosensor current followed rise in venous lactate but for participants 4 and 5, there was increase in biosensor current before measurable rises in venous lactate. We also observed different patterns of concordance between biosensor current and venous lactate for all participants between the exercise and rest phase: there was a greater lag in biosensor change with respect to venous lactate during the rest phase, suggesting a slower relative ISF lactate clearance in skin compared to blood.

**Figure 2.**
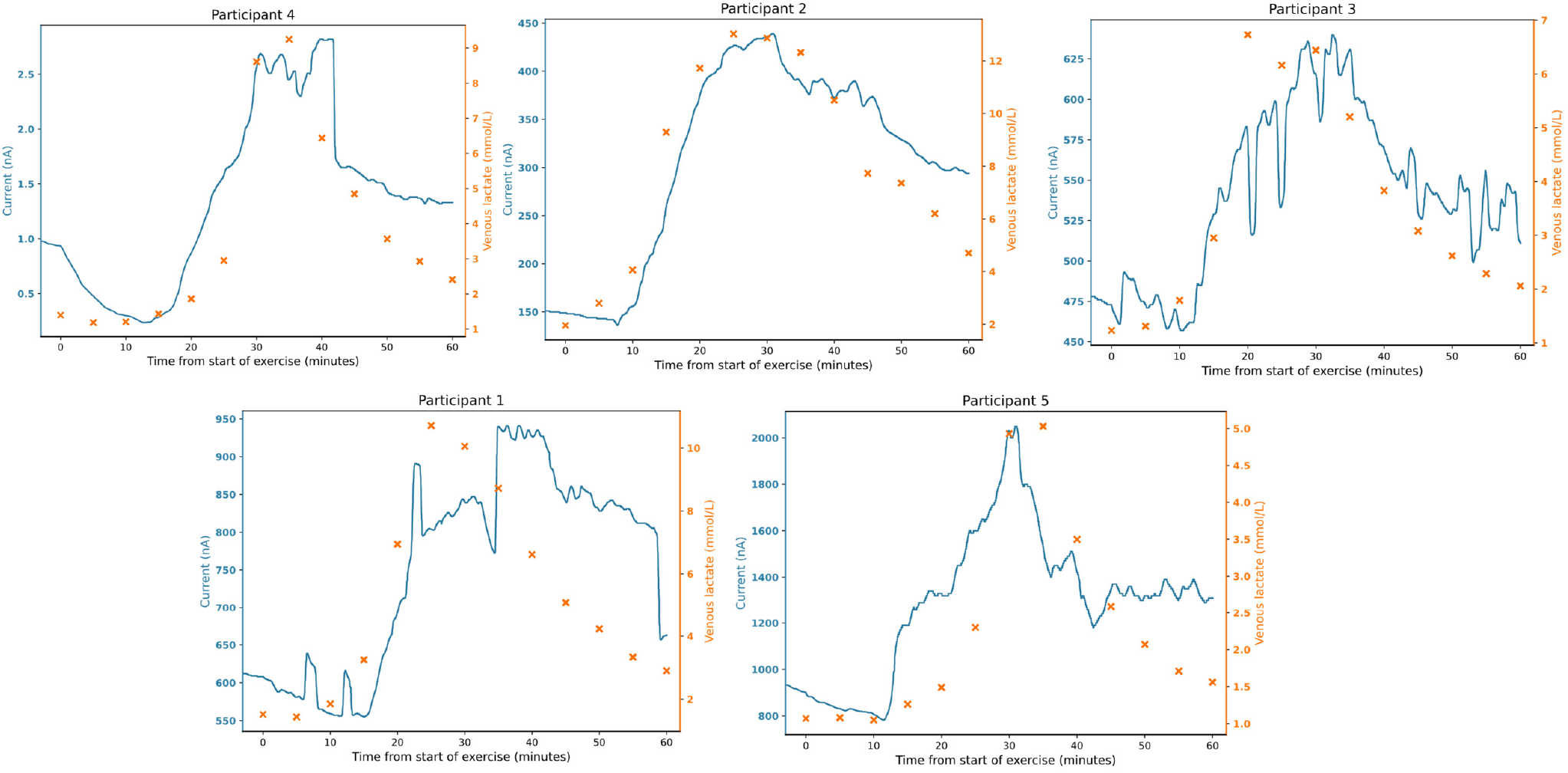
Microneedle biosensor current (blue continuous), venous lactate sampling (orange crosses) against time for individual participants (n=5). Exercise commenced at 0 minutes and stopped at 30 minutes, followed by a rest phase until the end of the study at 60 minutes.

Microdialysis was performed in participant 5 to understand trends in ISF lactate concentration, and to provide a reference measurement between biosensor current and blood lactate (Figure 3). The dialysate and venous lactate were in good agreement with a proportionate relationship up to 5 mmol/L during exercise and rest (Pearson’s r = 0.76). There was also a positive relationship between dialysate lactate concentration and biosensor current (Pearson’s r=0.58). We observed that ISF lactate concentration were significantly lower compared with that of blood. Dialysate lactate concentration is a function of probe recovery and determined by a number of factors including membrane length, perfusate flow rate and inherent differences between compartments. The results obtained therefore represents only a fraction of the total ISF concentration.

**Figure 3.**
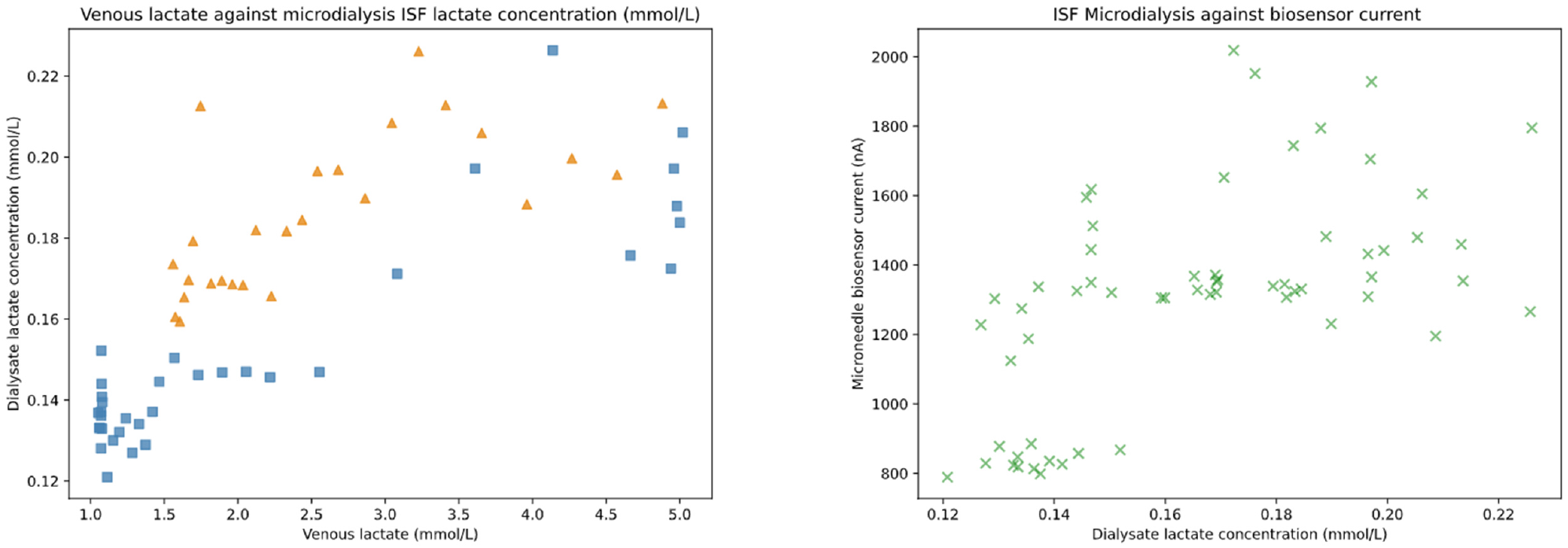
Left: Venous lactate plotted against dialysate lactate concentrations from microdialysis. Blue squares represent the exercise phase and the orange triangles represent the rest phase of the study. Right: Green crosses represent dialysate lactate concentrations against microneedle biosensor current downsampled to 1-minute intervals.

Venous blood lactate was used to calibrate biosensor current in order to compare agreement between measurements. We calibrated the biosensors separately depending on whether the data was obtained from the exercise, or rest phase given patterns of microneedle current:venous lactate relationships observed. In general, comparison in both phases show good overall mean agreement with a 95% confidence interval difference of +/- 1.89 mmol/L (figure 4).

**Figure 4.**
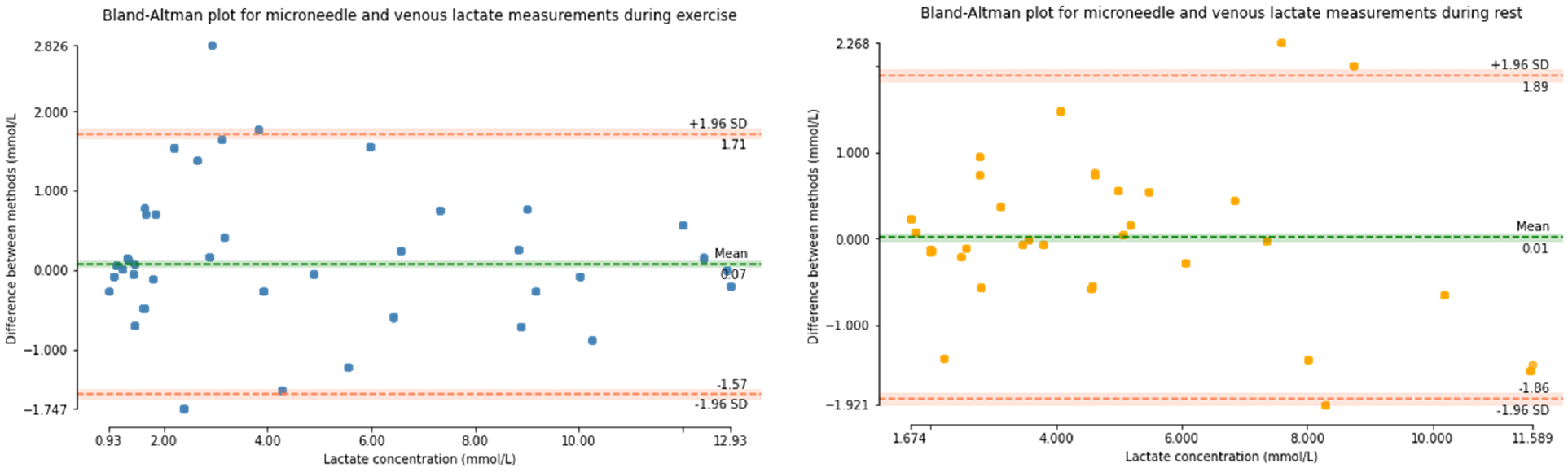
Bland-Altman plots of agreement between biosensor and venous blood lactate aggregated for all 5 participants. Analyses were separated by phase of the study between exercise and rest. The left plot shows data obtained during exercise (0-30 minutes), and the right plot shows data during rest (30-60 minutes). The horizontal axes show the range of lactate observed in the study and vertical axes difference in agreement between measurements. The 95% confidence interval (+/- 1.96 of standard deviation) is shown in orange horizontal lines.

We analysed performance of the biosensors to detect the change in lactate over time, given lactate clearance represented a clinically-relevant endpoint. Biosensor and venous lactate data were normalised to the same unit and dynamics represented by calculating the mean gradient of a rolling-window average spanning 5 minutes throughout the study (Figure 5). The dynamics of aggregated lactate change measured by the microneedle followed that of venous blood and with a Pearson’s coefficient of 0.86, showing correlation between measurements. The median response times estimated by differences in the peak measurements at the inflection point between biosensor current and interpolated lactate concentrations was 5.0 (IQR -4.0 to 11.0) minutes.

**Figure 5.**
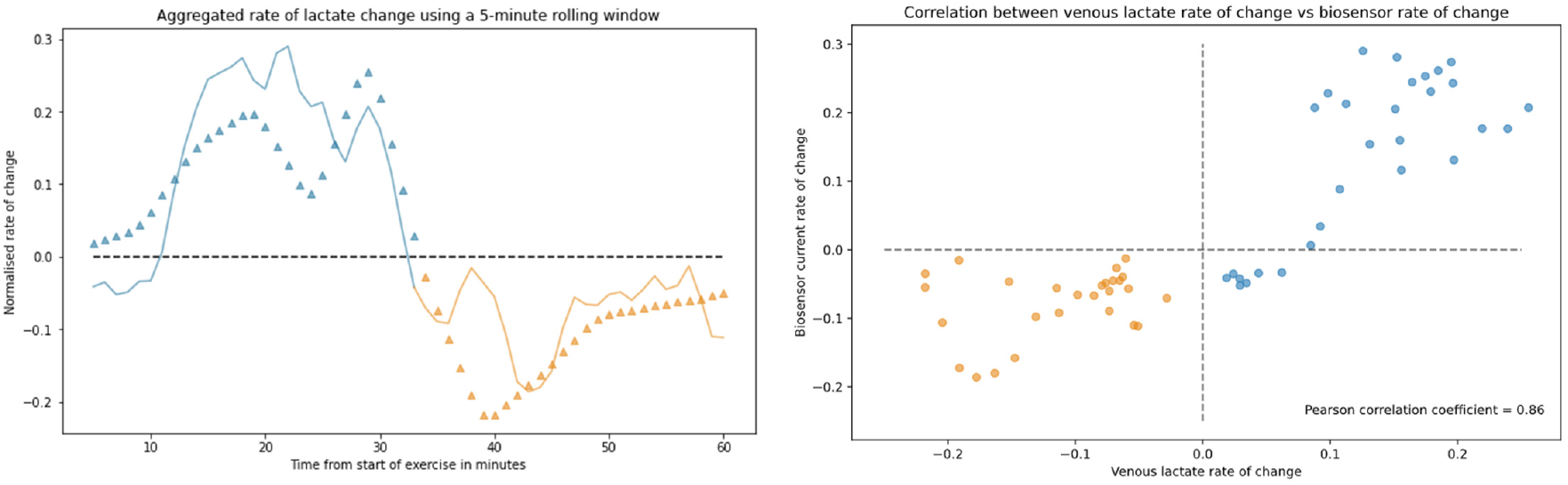
Left: normalised rate of change for all participants using a rolling window against time. The continuous line shows rate of change for the biosensor and triangles show venous lactate change at 1-minute intervals. The blue plots represent exercise phase and orange represents rest phase. The mean lactate peak occurred after 32 minutes after start of exercise. Right: Correlation scatterplot showing correlation between two measurements with a r of 0.86.

A visual analogue score in a questionnaire was administered to the participants at the end of the study after 2 hours of microneedle biosensor placement on the forearm. The mean score for discomfort at the site of the biosensor was 0.4/10, and the amount of restriction for the biosensor was rated 2.9/10 with comments relating to the wiring connecting the biosensor to the potentiostat. A photograph of the skin at 0, 15, 60 minutes on removal of the biosensor after 120 minutes of placement for an individual participant is shown in Figure 6 showing almost complete resolution of skin changes by 60 minutes of microneedle removal. No adverse events were reported during or after the study.

**Figure 6.**
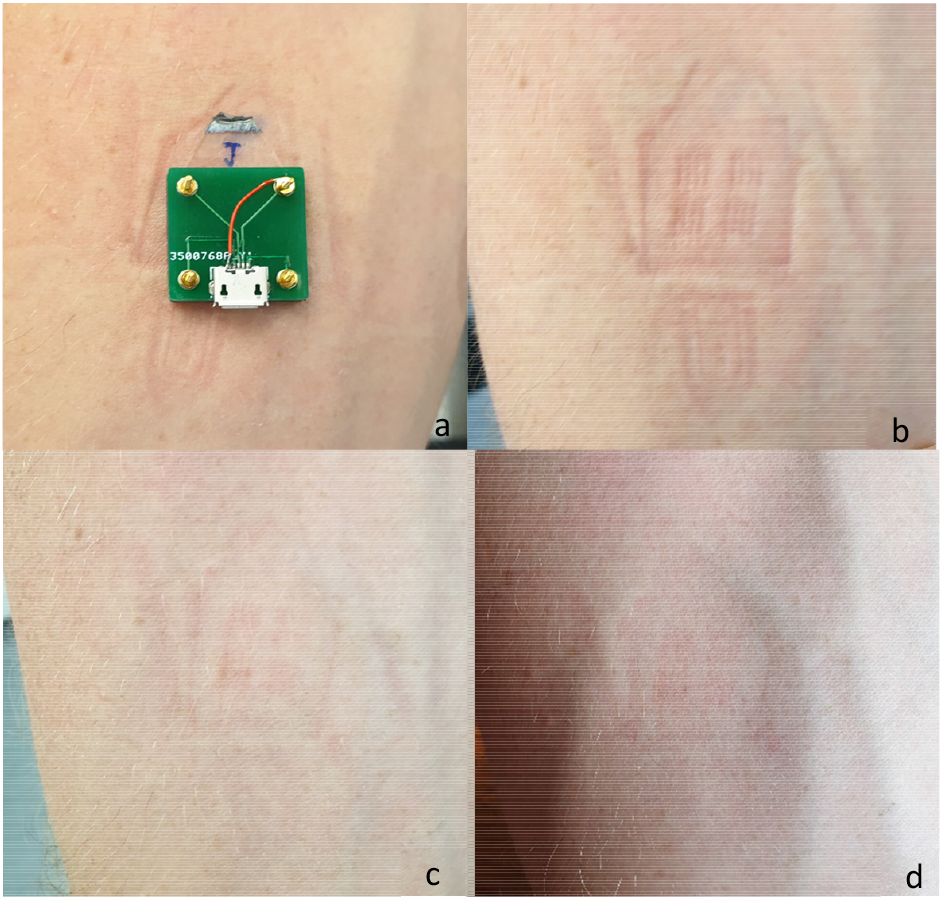
photographs showing: a) lactate biosensor in situ on participant forearm without connecting wires; b) underlying skin after immediate removal of biosensor; c) underlying skin after 15 minutes of biosensor removal; d) underlying skin after 60 minutes of biosensor removal

## Discussion

This is a first-in-human pilot validating performance of a minimally invasive lactate microneedle biosensor in healthy volunteers. The biosensor provides a self-contained modality for measuring ISF lactate continuously and in real-time. We show that the microneedle biosensor placed on the forearm was able to detect lactate generated from leg exercise. Venous lactate ranges between 1.07 to 13.03 mmol/L were measured and biosensor current increased on average within 5 minutes of a change in venous lactate, showing correlation in terms of both levels and dynamics over time. Consistent results from microneedle biosensor signals were seen for all participants during the study period and the biosensor was well tolerated for the 2-hour duration of use.

Lactate concentrations in pathological states is a sensitive, but non-specific biomarker. We used exercise as a proxy means to study transient increases in lactate. Although the physiological mechanisms for lactate production and clearance in exercise are different to that observed in pathology^3^, our model is valid in capturing the end manifestation of hyperlactataemia. Increases in microneedle biosensor current closely followed venous lactate generation during exercise but the biosensor exhibited a slower decrease during rest. This has been described previously in exercise^24^ and may relate to individual physiological variability as well as a relative reduction in perfusion to skin tissue and ISF compartments post-exercise, leading to delayed lactate clearance. In participants 4 and 5, we observed increases in microneedle current before that of venous lactate: possible explanations include biosensor and physiological variation, increased localised generation of lactate at the site placement and factors relating to the nature of microneedle placement within the skin.

Our microdialysis results support a relationship between ISF and venous lactate in exercise as well as microneedle current with ISF lactate concentrations. A more complete understanding of these relationships, as well as association between ISF lactate dynamics with clinical outcome will be a priority. In conditions such as sepsis, build-up of ISF lactate measured through microdialysis preceded changes in blood concentrations^15^. The measurement of ISF lactate could therefore provide clinically-actionable information prior to the onset of hyperlactatemia which is regarded as gold-standard. However, these relationships are not consistently seen across clinical settings^13^ nor is it clear how states of impaired microvascular perfusion, such as that seen in *falciparum* malaria^26^ affect ISF lactate dynamics.

In a previous study using the same microneedle platform, biosensors retained performance up to 24 hours of continuous use^20^. These are attractive characteristics for use in a range of clinical conditions, particularly for low and middle-income settings where a robust device without moving parts supports its implementation. The ease of insertion and patient acceptability are also major advantages for their use in settings with limited healthcare resources, and furthermore supports a role in research, allowing for detailed interrogation of physiology in settings of shock particularly in children^27^. The production of the device is scalable and the base cost is low^18^, supporting implementation across a variety of settings. Expansion of sensing modalities using this platform is possible, allowing for multi-modal detection of relevant substrates, other biomarkers or therapeutic indices^21^ – and will increase specificity of the tool. Linkage with decision-support systems and connectivity could provide benefits particularly in pre-hospital or ambulatory care settings^28^. Carefully-designed clinical studies will need to be carried out in order to investigate if continuous ISF lactate measurement ultimately translates into clinical benefit over intermittent blood measurements.

Limitations to our study include the small sample size and relatively short duration of biosensor use. Considerable sensor-to-sensor variation in terms of current output was also observed in light of small-scale fabrication processes and the pilot nature of the study. Subsequent design and testing iterations will also need to address performance of the biosensor particularly in the low perfusion states observed in clinical settings: derangements in local acid-base balance resulting from shock or hypoxia could result in markedly different relationships for lactate between ISF and blood. Placement of the biosensor, and the composition of underlying subcutaneous tissue as well as depth of insertion could play a role in individual variability in our study^29^. Standardised insertion methods onto the skin and methods of maintaining a consistent depth of penetration warrant investigation. Cross-reactivity with other compounds in ISF are known limitations for biosensors^30^. Substrates such as uric acid, ascorbic acid and paracetamol undergo redox reactions at potentials similar to that used in lactate sensing, and can result in an increase in biosensor current. Although the Nafion membrane used has been shown to protect against these interferents^31^and these compounds are not expected to change substantially during exercise, changes were observed changes on the control electrode (equivalent to the working sensor but without the enzyme). The implications of these changes are unclear and further work in clarifying the significance of these signals is ongoing within our group. Ensuring specificity in detection will be of importance for future use in clinical settings: improvements in biosensor designs, including the ability to reduce the potential applied at the electrode such as through use of direct electron transfer enzymes would offer significant benefits.

In conclusion, we demonstrate in a proof-of-concept study that the continuous measurement of ISF lactate using a minimally-invasive microneedle biosensor is feasible, well tolerated and produces clinically-actionable information. Work is ongoing to translate these findings into the clinical setting.

### Contributions

DKM and AHH conceived and designed the study. SJ, SANG, DMEF, AEGC, DOH were responsible for the design, fabrication and all technical aspects of the microneedle biosensor. SANG and MGB were responsible for microdialysis. DKM, RW and AHH performed the clinical study. DKM performed the analyses and wrote the first draft of the manuscript. All authors contributed to the revision of the manuscript and have approved the final version to be published.

## Data Availability

Data is available upon request to the corresponding author

## Acknowledgements

The research was funded by the Department of Health and Social Care, Centre for Antimicrobial Optimisation, at Imperial College, London. This report is independent research funded by the Department of Health and Social Care.

Infrastructure support was provided by the NIHR Imperial Biomedical Research Centre and the NIHR Imperial Clinical Research Facility.

AHH is a National Institute for Health Research (NIHR) Senior Investigator. DKM is supported by the Wellcome Trust [215010/Z/18/Z]

The views expressed in this publication are those of the author(s) and not necessarily those of the Department of Health and Social Care, NHS, or the National Institute for Health Research.

## Data availability

Data is available upon request to the corresponding author.

## References

1 Kruse O, Grunnet N, Barfod C. Blood lactate as a predictor for in-hospital mortality in patients admitted acutely to hospital: a systematic review. Scand J Trauma Resusc Emerg Med 2011; 19: 74.

2 Zhang Z, Xu X. Lactate clearance is a useful biomarker for the prediction of allcause mortality in critically Ill patients: A systematic review and meta-analysis. Critical Care Medicine 2014; 42: 2118–25.

3 Garcia-Alvarez M, Marik P, Bellomo R. Sepsis-associated hyperlactatemia. Crit Care 2014; 18: 503.

4 Singer M, Deutschman CS, Seymour CW, et al. The Third International Consensus Definitions for Sepsis and Septic Shock (Sepsis-3). JAMA 2016; 315: 801.

5 Aramburo A, Todd J, George EC, et al. Lactate clearance as a prognostic marker of mortality in severely ill febrile children in East Africa. BMC medicine 2018; 16: 37.

6 Yacoub S, Trung TH, Lam PK, et al. Cardio-haemodynamic assessment and venous lactate in severe dengue: Relationship with recurrent shock and respiratory distress. PLoS neglected tropical diseases 2017; 11: e0005740.

7 Odom SR, Howell MD, Silva GS, et al. Lactate clearance as a predictor of mortality in trauma patients. J Trauma Acute Care Surg 2013; 74: 999–1004.

8 Zymliński R, Biegus J, Sokolski M, et al. Increased blood lactate is prevalent and identifies poor prognosis in patients with acute heart failure without overt peripheral hypoperfusion. European Journal of Heart Failure 2018; 20: 1011–8.

9 Govender P, Tosh W, Burt C, Falter F. Evaluation of Increase in Intraoperative Lactate Level as a Predictor of Outcome in Adults After Cardiac Surgery. J Cardiothorac Vasc Anesth 2020; 34: 877–84.

10 Goodwin ML, Harris JE, Hernández A, Gladden LB. Blood Lactate Measurements and Analysis during Exercise: A Guide for Clinicians. J Diabetes Sci Technol 2007; 1: 558–69.

11 Graham CA, Leung LY, Lo RS, et al. Agreement between capillary and venous lactate in emergency department patients: prospective observational study. BMJ Open 2019; 9: e026109.

12 Wilson ML, Fleming KA, Kuti MA, Looi LM, Lago N, Ru K. Access to pathology and laboratory medicine services: a crucial gap. The Lancet 2018; 391: 1927–38.

13 Schierenbeck F, Nijsten MWN, Franco-Cereceda A, Liska J. Introducing intravascular microdialysis for continuous lactate monitoring in patients undergoing cardiac surgery: a prospective observational study. Critical Care 2014; 18: R56.

14 Cass AEG, Sharma S. Microneedle Enzyme Sensor Arrays for Continuous In Vivo Monitoring. Methods Enzymol 2017; 589: 413–27.

15 Kopterides P, Nikitas N, Vassiliadi D, et al. Microdialysis-assessed interstitium alterations during sepsis: Relationship to stage, infection, and pathogen. Intensive Care Medicine 2011; 37: 1756–64.

16 Ellmerer M, Haluzik M, Blaha J, et al. Clinical Evaluation of Subcutaneous Lactate Measurement in Patients after Major Cardiac Surgery. International Journal of Endocrinology 2009; 2009: e390975.

17 Kopterides P, Theodorakopoulou M, Ilias I, et al. Interrelationship between blood and tissue lactate in a general intensive care unit: A subcutaneous adipose tissue microdialysis study on 162 critically ill patients. Journal of Critical Care 2012; 27: 9–742.

18 Sharma S, Saeed A, Johnson C, Gadegaard N, Cass AE. Rapid, low cost prototyping of transdermal devices for personal healthcare monitoring. Sens Biosensing Res 2017; 13: 104–8.

19 Gill HS, Denson DD, Burris BA, Prausnitz MR. Effect of microneedle design on pain in human subjects. Clin J Pain 2008; 24: 585–94.

20 Sharma S, El-Laboudi A, Reddy M, et al. A pilot study in humans of microneedle sensor arrays for continuous glucose monitoring. Anal Methods 2018; 10: 2088–95.

21 Rawson TM, Gowers SAN, Freeman DME, et al. Microneedle biosensors for real-time, minimally invasive drug monitoring of phenoxymethylpenicillin: a first-in-human evaluation in healthy volunteers. The Lancet Digital Health 2019; 1: e335–43.

22 Rawson TM, Ming D, Gowers SA, et al. Public acceptability of computer-controlled antibiotic management: An exploration of automated dosing and opportunities for implementation. Journal of Infection 2018; published online Aug. DOI:10.1016/j.jinf.2018.08.005.

23 Bollella P, Sharma S, Cass AEG, Antiochia R. Microneedle-based biosensor for minimally-invasive lactate detection. Biosens Bioelectron 2019; 123: 152–9.

24 Gowers SAN, Curto VF, Seneci CA, et al. 3D Printed Microfluidic Device with Integrated Biosensors for Online Analysis of Subcutaneous Human Microdialysate. Anal Chem 2015; 87: 7763–70.

25 Acciaroli G, Vettoretti M, Facchinetti A, Sparacino G. Calibration of Minimally Invasive Continuous Glucose Monitoring Sensors: State-of-The-Art and Current Perspectives. Biosensors (Basel) 2018; 8: 24.

26 Hanson J, Lee SJ, Hossain MA, et al. Microvascular obstruction and endothelial activation are independently associated with the clinical manifestations of severe falciparum malaria in adults: an observational study. BMC Medicine 2015; 13: 122.

27 Maitland K, George EC, Evans JA, et al. Exploring mechanisms of excess mortality with early fluid resuscitation: insights from the FEAST trial. BMC medicine 2013; 11: 68.

28 Ming D, Rawson T, Sangkaew S, Rodriguez-Manzano J, Georgiou P, Holmes A. Connectivity of rapid-testing diagnostics and surveillance of infectious diseases. Bull World Health Organ 2019; 97: 242–4.

29 Enderle B, Moser I, Kannan C, Schwab KO, Urban G. Interstitial Glucose and Lactate Levels Are Inversely Correlated With the Body Mass Index: Need for In Vivo Calibration of Glucose Sensor Results With Blood Values in Obese Patients. J Diabetes Sci Technol 2018; 12: 341–8.

30 Sasso SV, Pierce RJ, Walla Robert, Yacynych AM. Electropolymerized 1,2-diaminobenzene as a means to prevent interferences and fouling and to stabilize immobilized enzyme in electrochemical biosensors. Anal Chem 1990; 62: 1111–7.

31 Romero MR, Ahumada F, Garay F, Baruzzi AM. Amperometric Biosensor for Direct Blood Lactate Detection. Anal Chem 2010; 82: 5568–72.

