## Supplementary appendix for "Real-time Continuous Measurement of Lactate through a Minimally-invasive Microneedle Biosensor: a Phase I Clinical Study"

Supplementary appendix – study protocol

**Study protocol**

Inclusion criteria

1. Consenting adults ≥ 18 years old
2. Healthy with no previously diagnosed medical condition from a medical practice or active medications
3. Able to perform moderately intensive exercise on an exercise bicycle without difficulty for at least 30 minutes continuously, and engages in regular aerobic exercise at least twice a week

Exclusion criteria

1. Active inflammatory skin condition such as eczema or dermatitis
2. Active soft tissue infection or infection at any site
3. Known hypersensitivity to any microneedle component or dressings
4. Presence of any implantable electronic devices such as a pacemaker or stimulators
5. Currently pregnant

**Study Method**

The study will take place in a research room within the NIHR-Imperial Clinical Research Facility. After screening and informed consent, the study will begin:

1) A lactate microneedle biosensor will be placed on the non-dominant forearm prepared by chlorhexidine/alcohol and connected to a potentiostat, which records data. There will be a run-in stabilisation time of approximately 60 minutes to normalise readings.
2) EMLA 5% topical anaesthetic cream will be applied to the site of microdialysis insertion.
3) A peripheral cannula will be inserted in the arm for blood lactate testing.
4) A microdialysis catheter will be inserted on the non-dominant forearm for the measurement of interstitial fluid lactate (see section below for description).
5) At 60 minutes into the study, baseline blood lactate and ISF lactate will be taken from the peripheral cannula and microdialysis catheter respectively.
6) The participant will be asked to begin exercise on an exercise bicycle starting at 90W, with increments in power every 5 minutes up to a maximum of 210W.
7) During the study, 2 ml of blood will be drawn from the peripheral cannula every 5 minutes for venous lactate determination, and will continue until the end of the rest period.
8) Microdialysis samples will also be drawn from the microdialysis catheter every 5 minutes for ISF lactate determination and will continue until the end of the study.
9) The exercise period will stop 90 minutes into the study (after 30 minutes of total exercise) and the rest period will begin for 30 minutes.
10) At the end of the rest period, all monitoring devices (microneedle, peripheral cannula, microdialysis catheter) will be removed from the participant.
11) The participant will be given a questionnaire to complete. The site of the microneedle placement on the skin will also be photographed by the study camera – this will not contain any participant identifiable information and will be consented for explicitly. The photograph will be stored electronically on servers maintained by Imperial College Healthcare NHS Trust within the NIHR-Imperial Clinical Research Facility.
12) End of study, and participant free to leave.
